# Explainable machine learning reveals the challenges of predicting new-onset motor complications in Parkinson’s disease

**DOI:** 10.64898/2026.07.06.26357357

**Authors:** Walter Endrizzi, Nicole Campese, Flavio Ragni, Monica Moroni, Stefano Bovo, Chiara Longo, Lorenzo Gios, Antonio Uccelli, Bruno Giometto, Giuseppe Jurman, Venet Osmani, Maria Chiara Malaguti, the NeuroArtP3 Network

**Author notes:** Corresponding Author mail address. NeuroArtP3 Network details are presented in the Author Information section.

## Abstract

Predicting new-onset motor fluctuations and levodopa-induced dyskinesias (LID) is crucial for optimizing Parkinson’s disease management. To establish a transparent prognostic framework, we applied explainable machine learning to real-world, multicentric clinical data from 247 patients to forecast the 3-year onset of these complications. Evaluated strictly on complication-free patients, the models achieved moderate predictive power (LID MCC=0.28; fluctuations MCC=0.32). SHAP-based interpretability confirmed predictions aligned accurately with established clinical knowledge, driven primarily by levodopa duration and Levodopa Equivalent Daily Dose, with risk increasing significantly above a 300-400 mg threshold. Crucially, an ablation study revealed that excluding patients with pre-existing complications from training caused model sensitivity to collapse, demonstrating that the full spectrum of disease severity is essential for robust risk stratification. Ultimately, this rigorous methodological stress-test demonstrates that baseline clinical features alone yield limited absolute sensitivity, highlighting the necessity of integrating dynamic, longitudinal data to achieve clinical-grade individualized prediction.

## Introduction

Motor complications are common, troublesome features associated with long-term Levodopa treatment in Parkinson’s disease (PD) (1). According to a longitudinal population-based study, up to 50% of persons living with PD experience motor fluctuations and 15% of PD patients develop Levodopa induced dyskinesias (LID) after 5 years of Levodopa treatment, with an incidence increasing to 100% and 55% respectively after 10 years (2). On the other hand, some people with PD do not experience any motor fluctuations, even after many years of Levodopa treatment, which defines the so called long-duration response to Levodopa (3). Several risk factors for motor complications have been identified in PD, with longer disease duration and higher total Levodopa Equivalent Daily Dose (LEDD) emerging as the strongest predictors of both motor fluctuations and LID (1,4). Additionally, female gender and a younger age at disease onset were consistently reported across studies as risk factors for both motor fluctuations and LID (5,6).

Predicting the onset of motor complications in patients with PD is crucial for tailoring patients’ care, improving their quality of life, preventing disability and identifying patients who may be eligible for device-assisted therapies (7,8).

To date, evidence on risk factors for motor fluctuations and LID relies primarily on single-cohort studies, limiting the generalizability of findings, and no validated algorithms are currently available to accurately predict their onset. To address these prognostic challenges, machine learning (ML) models are being increasingly explored, often in combination with Explainable Artificial Intelligence (XAI) approaches that can help interpret learned predictive patterns and assess their clinical plausibility (9–11). While XAI may also support the generation of new biological or clinical hypotheses, its role is particularly important in predictive modelling to determine whether model decisions are consistent with established disease knowledge rather than driven by spurious associations. However, translating these promising tools into practice requires evaluations that demonstrate genuine clinical utility. Rather than relying solely on broad dataset metrics, rigorous assessments should directly test the true clinical challenge: predicting the future onset of motor complications in patients currently free of them.

This study utilizes real-world clinical data from two distinct cohorts to develop and evaluate ML models for predicting the onset of motor fluctuations and LID in persons with PD within three years from baseline assessment.

## Results

### LID Prediction

Model evaluation for future LID prediction was primarily based on the MCC, a metric chosen for its robustness in imbalanced classification (prevalence: 39% total, 31% evaluation subset). While the highest MCC scores were generally achieved when training and testing on the full cohort (SVC MCC = 0.46 ± 0.11; XGB MCC = 0.42 ± 0.11), these performances were considered inflated by the inclusion in the evaluation set of patients with LID at baseline. Therefore, to maximize clinical relevance, final model selection was based on performance specifically within the subset of patients who were free from LID at baseline. The best-performing models in this clinically relevant analysis were the SVC and XGB, which were selected for detailed evaluation (see Supplementary Table S3 for all performance metrics).

Focusing on the task of predicting only the new onset of LID, the two models achieved moderate predictive power (SVC MCC 0.28 ± 0.14; XGB MCC 0.26 ± 0.16). However, when the training strategy was altered to entirely exclude patients free from LID at baseline, a severe performance drop was observed, particularly for the tree-based model (XGB MCC 0.02 ± 0.11). To further evaluate the clinical reliability of these predictions, Positive Predictive Value (PPV) and Negative Predictive Value (NPV) were systematically assessed. In the clinically relevant subset, the SVC model achieved a PPV of 0.55 ± 0.16 and an NPV of 0.78 ± 0.03, while the XGB model yielded a PPV of 0.48 ± 0.15 and an NPV of 0.79 ± 0.04 (Supplementary Table S3).

To understand the key drivers behind the predictions of the SVC model, a SHAP analysis was conducted. The global feature importance is visualized in the summary plot (Figure 1A), which reveals that LEDD, motor fluctuations at baseline, and duration of Levodopa therapy were the three most influential features for SVC in predicting LID. For all three features, larger values (indicated by red dots) corresponded to positive SHAP values, indicating a strong contribution toward predicting the presence of future LID. Conversely, low values for these features (blue dots) were associated with negative SHAP values, driving the model to predict absence of LID within three years from the baseline.

**Figure 1.**
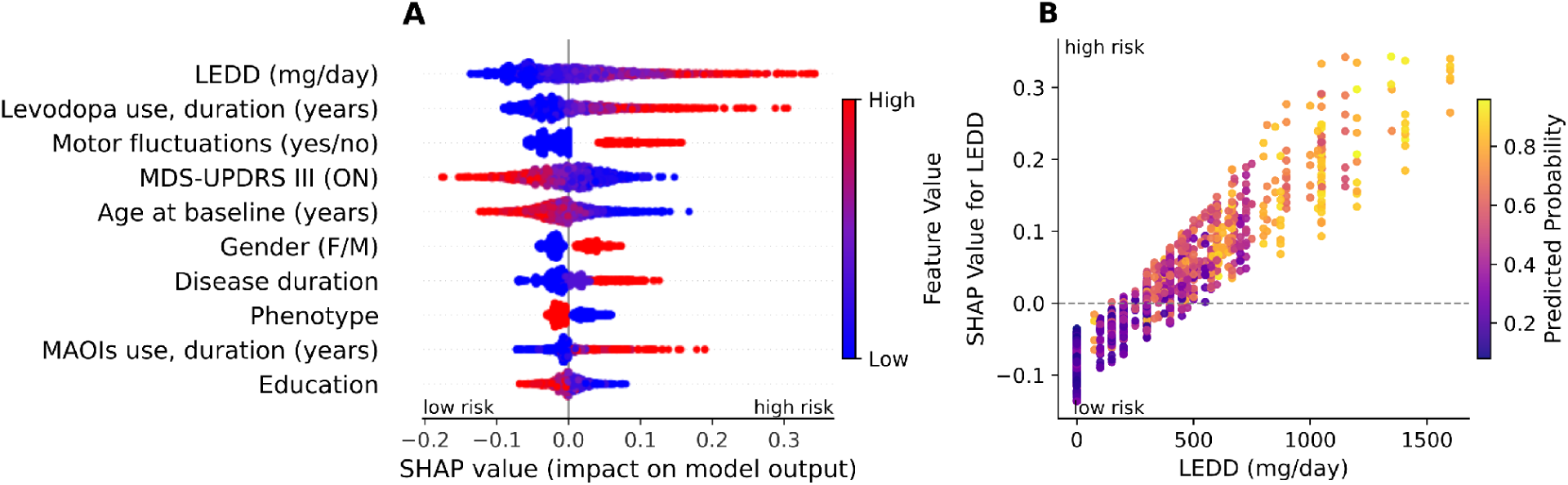
SHAP analysis of SVC for the prediction of future LID. **A.** Global feature importance and impact for the SVC model revealed by SHAP. Features are ranked in descending order of importance. Each dot represents a patient from the test set. The horizontal axis shows the SHAP value, indicating the feature’s contribution to predicting LID (positive values) or absence of LID (negative values) within the third year. Color indicates the feature’s original value (blue=low, red=high), revealing the direction and magnitude of its effect. **B.** Detailed analysis of the LEDD feature using a SHAP dependence plot for the SVC model. The x-axis plots the patient’s actual LEDD on its original scale, while the y-axis plots the SHAP value, indicating the feature’s contribution to the prediction. A potential dosage threshold is visible around 300-400 mg, where the feature’s impact (SHAP value) transitions from negative (low risk) to positive (high risk). LEDD: Levodopa equivalent daily dose; LID: Levodopa-induced dyskinesia; MAOIs: monoamine oxidase inhibitors.

Given its high importance and clinical relevance, the LEDD feature was analyzed in greater detail to investigate whether a specific dosage threshold is associated with high risk of future LID. The SHAP dependence plot (Figure 1B) revealed that doses below approximately 300-400 mg cluster around or below a SHAP value of zero, suggesting a minimal impact on the model’s prediction. In contrast, at doses above this 300-400 mg threshold, the SHAP values become almost exclusively positive, indicating that higher Levodopa doses significantly increase the model’s predicted risk of developing LID.

This interpretability analysis was also performed on the XGB model. The results were highly consistent with SVC: the SHAP analysis identified the same key drivers of risk, and the LEDD dependence analysis revealed an identical 300-400 mg risk threshold. The full SHAP summary and dependence plots for the XGB model are provided in the Appendix (Figure S2).

### Motor fluctuations prediction

For the prediction of motor fluctuations, across all tested classifiers (Supplementary Table S4), the Voting ensemble and RF were the best performing models and were selected for further analysis. Consistent with the LID task, performance metrics were significantly inflated when evaluated on the full cohort (Table S4, MCC Voting = 0.51 ± 0.12, MCC RF = 0.53 ± 0.11). Focusing on the clinically relevant prediction of new onset in patients free of motor complications at baseline, a substantial performance drop was observed for both models (Voting MCC = 0.32 ± 0.18, RF MCC = 0.32 ± 0.18).

When models were trained directly on patients without baseline motor fluctuations, a further performance decline was observed (Voting MCC = 0.08 ± 0.15, RF MCC = 0.10 ± 0.15), mirroring the trend observed in the LID analysis. Similarly, the assessment of predictive values highlighted the models’ conservative thresholding behavior. When evaluated directly on patients without baseline motor fluctuations, both the Voting ensemble and the RF model yielded a PPV of 0.51 ± 0.13 and an NPV of 0.81 ± 0.06 (Supplementary Table S4).

To identify the key drivers for the motor fluctuations prediction, a SHAP analysis on the best performing models was performed. The resulting global feature importance for the Voting ensemble model is shown in the summary plot (Figure 2), which reveals a SHAP pattern consistent with that of the RF model (Figure S4).

**Figure 2.**
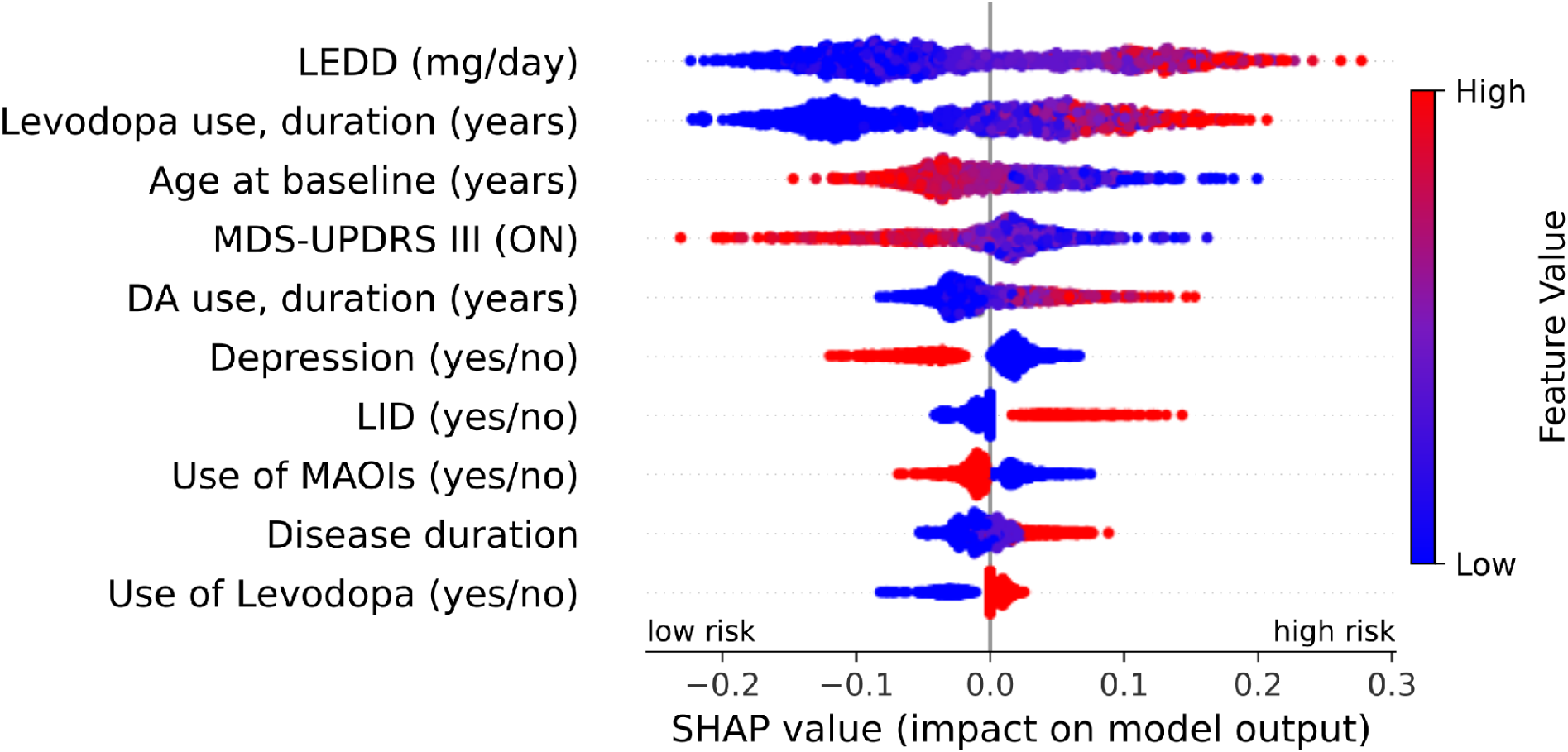
SHAP analysis of Voting ensemble for prediction of future motor fluctuations. Global feature importance and impact for the Voting ensemble revealed by SHAP. Features are ranked in descending order of importance. Each dot represents a patient from the test set. The horizontal axis shows the SHAP value, indicating the feature’s contribution to predicting the presence (positive values) or absence of motor fluctuations (negative values) within the third year. Colors indicate the feature’s original value (blue=low, red=high), revealing the direction and magnitude of its effect. DA: dopamine agonist; LEDD: levodopa equivalent daily dose; LID: Levodopa-induced dyskinesia; MAOIs: monoamine oxidase inhibitors.

### Impact of Training Cohort Composition

A controlled ablation study was performed for both clinical tasks to investigate the performance collapse observed when training exclusively on patients without motor complications at baseline. First, the effect of sample size was assessed. When a random subset of patients matched in size to those with baseline complications was removed from training (Supplementary Tables S3, S4), predictive performance remained robust and comparable to full-cohort training for both outcomes. This indicates that the observed deterioration was not driven by reduced cohort size, but by the loss of specific clinical information.

Second, model evaluation revealed a dissociation between discrimination and threshold-based decision performance. Rank-based metrics were relatively preserved (e.g., SVC AUC-ROC for LID: 0.67 ± 0.08 vs 0.70 ± 0.09 after random removal), whereas decision-based metrics collapsed, with MCC falling to ∼0.10 and sensitivity decreasing by more than 50%. Thus, models could still rank patients by risk but failed to assign calibrated probabilities high enough to cross the decision threshold.

Across architectures, boosting and randomized ensembles showed the strongest collapse, whereas RF and linear models were somewhat more stable. However, sensitivity fell below 0.25 across all models, indicating that no algorithm was unaffected by the exclusion of patients with pre-existing motor complications.

## Discussion

Consistently with previous studies (4), a longer cumulative exposure to Levodopa, a higher LEDD and the presence of motor fluctuations at baseline were the strongest determinants of LID at follow-up in our cohort. Moreover, a longer disease duration at baseline was associated with a higher subsequent risk of LID. These results are consistent with observations by Cilia and colleagues (4), showing that the disease duration and the cumulative duration of the exposure to Levodopa, rather than the disease duration at the time of Levodopa initiation, are the stronger predictors of both motor fluctuations and LID in PD. A higher LEDD has been consistently reported as a risk factor for the development of LID, with critical LEDD thresholds lying between 365 and 400 mg daily, according to different studies (4,19). This association was confirmed by our model, which showed a substantial increase of the risk of LID for LEDD values greater than 300-400 mg daily, in line with previous evidence and supporting the clinical plausibility of the learned predictive pattern. Paralleling previous observations (19–21), female PD patients were found to be at higher risk of LID. Both lower body weight and gender-related differences in the pharmacokinetics of Levodopa may underlie this finding (22). Similarly, previous observations on the association between younger age at disease onset and increased LID risk were confirmed (4,19,23–25). In young-onset PD, pre- and post-synaptic neuronal plasticity mechanisms are supposed to effectively compensate the dopaminergic neuronal loss until the very late stages of the disease, in which a more severe depletion of dopaminergic cells occurs (4,26,27). Young-onset PD patients are therefore supposed to present with a more severe dopaminergic loss compared to older-onset persons with PD with similar severity of motor features, thus accounting for an increased risk of motor fluctuations (4,26).

In our cohort, higher MDS-UPDRS III scores at baseline were associated with a lower subsequent risk of motor fluctuations. This is not consistent with what reported by a previous study by Eusebi and colleagues (20). It is worth noting that the tremor items have a greater relative weight in determining the total score MDS-UPDRS III compared to akinesia and rigidity items. Therefore, persons presenting a tremor-dominant phenotype tend to score higher at the MDS-UPRDS III compared to rigid-akinetic phenotypes. In our cohort, PD patients with a tremor-dominant phenotype show a lower risk of future LID. The association observed between MDS-UPDRS III scores, and the risk of LID may be therefore driven by differences in clinical phenotypes rather than in disease severity.

In our model, motor fluctuations at follow-up were associated with a higher LEDD, a longer cumulative exposure to Levodopa, and a younger age at baseline. Similarly to LID, both the higher LEDD, a longer cumulative exposure to Levodopa and a longer disease duration, are well established risk factors for motor fluctuations in PD (4,21). In our cohort, motor fluctuations were associated with lower MDS-UPDRS III scores. Prior studies report conflicting findings, showing either a positive (2,5,28) or no association between MDS-UPDRS III scores (21). As previously speculated for LID, the higher MDS-UPDRS III motor scores at baseline may reflect differences in the clinical phenotype rather than in disease severity alone and therefore justify our observation. Depression emerged as a protective factor for motor fluctuations in our models.

Non-dopaminergic pathways, including serotoninergic and noradrenergic pathways, seem to be involved in the pathophysiology of motor fluctuations in PD. Persons with depression may be treated with drugs acting on these neurotransmitters, which may therefore exert an indirect effect on motor fluctuations (29). In our cohort the presence of LID at baseline was associated with an increased risk of subsequent motor fluctuations. Motor fluctuations and LID are strongly associated phenomena that frequently co-occur in advanced stage of PD (30). From a pathophysiological perspective, shared pathological changes affecting the basal ganglia dopaminergic networks may favor both motor fluctuations and LID. In fact, as the nigrostriatal denervation becomes more severe with disease progression, changes in the dopaminergic receptor sensitivity to Levodopa occur, making PD patients more dependent on the pulsatile oral administration of the drug. This may lead to motor fluctuations with sudden transitions from “on” and “off” states, which are typically counteracted by an increase of the oral Levodopa intake that may in turn favor the development of LID (4,26).

A critical finding of our ablation study was that the inclusion of patients with pre-existing motor complications in the training set appears to be important for model sensitivity, even when the clinical aim is to predict new onset in patients currently free of these complications.

The dissociation observed between preserved discrimination (AUC-ROC, which reflects the model’s intrinsic ranking ability independent of threshold) and collapsed decision-making (MCC) highlights an important limitation of the modelling framework. The random patient removal control demonstrated that the collapse in threshold-dependent metrics was not an artifact of reduced training sample size. While the models maintained stable rank-based discrimination after excluding baseline-complicated patients (e.g., SVC AUC-ROC for LID at 0.67 ± 0.08, compared to 0.70 ± 0.09 in the size-matched control), they lost the ability to confidently assign high absolute probabilities. The cohort without baseline motor complications provided sufficient information for the models to learn the relative risk ranking but lacked the examples necessary to scale these predictions into high probabilities.

In the absence of patients with evident motor complications acting as high-confidence reference cases, the models learned a compressed probability distribution. This uncertainty was partly driven by an artificially lowered baseline prevalence; excluding prevalent cases skewed the study cohort and reduced the overall probability of a positive outcome. To account for this lower prevalence and the subtler clinical presentation of newly incident cases, models adjusted their overall risk estimates downwards. Consequently, even the highest-risk patients received conservative scores that rarely crossed the clinical threshold for detection.

This suggests that patients with pre-existing complications serve as anchor examples, establishing the upper bound of the risk probability scale, rather than acting as a substitute for learning early predictive features. Without these extremes the model lacks a reference for high-confidence prediction. This dependency was observed consistently across model architectures: no single algorithm demonstrated immunity to the loss of these reference cases. These results support the intuition that to confidently detect the subtle signs of onset in patients free from motor complications, model training benefits from including the full spectrum of disease severity, even advanced cases, to establish a robust probabilistic frame of reference.

Crucially, while our ablation study demonstrates that including the full spectrum of disease severity is mathematically essential to maximize model sensitivity, our stress-testing approach also highlights the current limits of clinical translation. When evaluating the true clinical challenge (i.e. predicting new-onset complications in patients currently free of them) the optimized models still miss a substantial proportion of incident cases (e.g., SVC sensitivity for LID of 0.37 ± 0.12). As we note, these models achieve moderate predictive power, but deploying a prognostic tool with this missing rate as a standalone screening device would provide dangerous false reassurance to patients and clinicians. This performance ceiling suggests a fundamental biological reality: it is highly unlikely that a single, cross-sectional snapshot of baseline clinical features is sufficient to fully encapsulate the complex, three-year longitudinal trajectory of motor complications. Achieving highly sensitive, individualized prediction will require moving beyond baseline features to incorporate dynamic, time-series clinical data, genetic profiles, or continuous objective monitoring.

Identifying individuals at risk of motor complications is crucial to improve quality of life, prevent disability, and support personalized management in PD (31). Although several risk factors for LID and motor fluctuations are well established, a systematic tool for risk stratification is still lacking, and previous ML studies in PD have remained largely exploratory (9).

In this multicentric longitudinal cohort study, this gap was addressed by introducing an ML workflow to develop and stress-test predictive classifiers for early LID and motor fluctuations. The models rely on routinely collected clinical variables widely available in neurological practice, without requiring advanced imaging, genetic, or biomarker data, thereby supporting scalability and real-world applicability across centers with varying technological resources. The workflow also emphasizes clinical utility by explicitly testing for instability and miscalibration. Combined with SHAP-based explainability and evaluation on a clinically relevant test population, this provides a transparent and unbiased assessment of future risk without inflating predictive performance. Therefore, the core strength of this work is providing a transparent framework for evaluating the real-world clinical utility of explainable machine learning models, demonstrating how clinically interpretable prediction workflows can both reproduce established knowledge and reveal the current limitations of baseline clinical data for individualized prognostic prediction.

The current study has some limitations. First, rigorous harmonization of two prospectively collected datasets reduced the effective sample size. In addition, international heterogeneity in treatment practices, drug availability, and socio-economic factors may influence motor outcomes, requiring validation in larger and more diverse cohorts. Genetic data were excluded because of limited availability. Moreover, baseline body weight was frequently missing, preventing the calculation of weight-adjusted LEDD, and gastrointestinal dysfunction was modeled within a broader autonomic symptom variable, potentially masking its specific contribution to motor complications. In addition, an inherent limitation of this study, and of similar prognostic modeling efforts, is the reliance on baseline clinical features to predict a dynamic multi-year trajectory. As evidenced by the models’ low absolute sensitivity, this approach risks providing false reassurance if used to rule out future complications, underscoring the necessity for future predictive models to integrate dynamic, longitudinal biomarker tracking. Future studies in larger cohorts should also investigate whether the methodological limitations observed here, particularly the dissociation between preserved risk ranking and poor threshold-based classification, extend to other motor and non-motor complications of PD, such as falls, freezing of gait, dystonia, impulse control disorder, or apathy. Finally, the follow-up period was limited to three years, in accordance with the harmonized NeuroArtP3 protocol. Although this ensured comparable observation windows across cohorts, longer follow-up may capture additional incident cases of motor fluctuations and LID and improve the assessment of long-term predictive performance.

## Methods

### Ethics statement and protocol

The study was approved by the Local Ethics Committees (Ethics Committee of Azienda Provinciale per i Servizi Sanitari of Trento and Ethics Committee of Liguria Region—Protocol number NET-2018-12366666-3-PD). The protocol for this study was published early in 2024 and refers to the NeuroArtP3 (NET-2018-12366666) project (12). Written informed consent was obtained from every participant, and all assessments were performed according to national and international guidelines.

### Cohort characteristics

A total of 256 patients with PD diagnosed according to current criteria (13) were recruited from the NeuroArtP3 project at two Northern Italian sites: Azienda Sanitaria Universitaria Integrata del Trentino (formerly APSS - Provincial Health Services, Trento, n=126) and IRCCS San Martino Hospital (HSM, Genoa, n=130). The final analytic cohort consisted of 247 subjects after exclusion of seven patients treated with Deep Brain Stimulation (DBS) or other advanced therapies and two patients with incomplete clinical information.

For each participant, data was collected longitudinally during routine clinical practice for three consecutive years. Data collection across centers followed the harmonized NeuroArtP3 protocol, which defined a common set of clinical variables, shared follow-up timepoints, and standardized data-entry procedures to ensure comparability between cohorts (12). For the predictive modeling, only variables collected at the first visit (i.e. baseline visit) were used as input features. These baseline features encompassed clinical-demographic features, including clinical rating scales (e.g., the Movement Disorders Society Unified Parkinson’s Disease Rating Scale part III subscores - MDS-UPDRS III, Hoehn & Yahr – H&Y), PD core motor and non-motor features (e.g., presence of impulse control disorder - ICD, sleep disorders, cognitive status), and medication history. A complete list of these variables and their characteristics is provided in Table 1.

**Table 1.** Characteristics of the study cohort (N=247). Categorical variables are presented as count (percentage). Continuous variables are presented as median (Q1, Q3). In case of missing data, the sample size for the variable of interest is reported in brackets. If no specific sample size is indicated, the variable was available for the full cohort. Legend: DA: dopamine agonist; H&Y: Hoehn and Yahr stage; ICD: impulse control disorder; LEDD: levodopa equivalent daily dose; LID: Levodopa-induced dyskinesia; MAOIs: monoamine oxidase inhibitors.

| Variable | Level | Count (%) |
| --- | --- | --- |
| Gender (Male = 0; Female=1) | Female | 96 (38.9%) |
| Disease duration | 0-5 years | 145 (58.7%) |
|  | 5-10 years | 63 (25.5%) |
|  | > 10 years | 39 (15.8%) |
| Family history (for PD) | Yes | 80 (32.4%) |
| H&Y stage | 1 | 77 (31.2%) |
|  | 2 | 128 (51.8%) |
|  | >2 | 42 (17.0%) |
| Motor phenotype | Tremor-dominant | 145 (58.7%) |
|  | Akinetic-rigid | 102 (41.3%) |
| Falls | Yes | 31 (12.6%) |
| Freezing of gait | Yes | 31 (12.6%) |
| LID | Yes | 34 (13.8%) |
| Motor fluctuations | Yes | 59 (23.9%) |
| Presence of dysautonomia | Orthostatic hypotension | 20 (8.1%) |
|  | Other domains | 68 (27.5%) |
|  | None | 159 (64.4%) |
| Cognitive status (n=242) | Normal | 217 (87.9%) |
|  | MCI | 19 (7.7%) |
|  | Dementia | 6 (2.4%) |
| Anxiety | Yes | 61 (24.7%) |
| Depression | Yes | 70 (28.3%) |
| ICD | Yes | 20 (8.1%) |
| Hyposmia (n=211) | Yes | 74 (30.0%) |
| Sleep/Wake disorders | Yes | 101 (41.3%) |
| Use of Levodopa | Yes | 172 (69.6%) |
| Use of DA | Yes | 173 (70.0%) |
| Use of MAOIs | Yes | 147 (59.5%) |
| LID at follow-up | Yes | 96 (38.9%) |
| Motor fluctuations at follow-up | Yes | 113 (45.7%) |
| **Attribute (n)** | **Median (Q1, Q3)** |  |
| Age at baseline (n=247) | 69 (63, 74) |  |
| Education (n=244) | 11 (8, 13) |  |
| Time between symptoms onset and diagnosis (years) (n=238) | 1 (0, 2) |  |
| MDS-UPDRS III (ON) (n=237) | 17 (11, 27) |  |
| LEDD (n=246) | 300 (0, 450) |  |
| Levodopa use, duration (years, n=240) | 2 (0, 5) |  |
| DA use, duration (years, n=244) | 1 (0, 4) |  |
| MAOIs use, duration (years, n=245) | 0 (0, 1) |  |

The binary outcome labels, indicating whether a patient developed LID or motor fluctuations within three years from the baseline visit were created using the follow-up data. LID were defined as the sudden peak-of-dose emergence of involuntary movement disorders and motor fluctuations as the sudden transition between functional “on” and “off” states (including wearing-off, delayed-on, no-on, unpredictable off phenomena) (8,14), as assessed by interview during the follow-up visits and reported in the medical charts. Descriptive statistics of these two outcomes are reported in Table 1. A crucial stratification was necessary for this analysis: at baseline, a number of patients already presented with LID (n=34) or motor fluctuations (n=59). The true clinical challenge lies in identifying which patients will develop these symptoms in the future, not in confirming a pre-existing condition. Therefore, to build a model with real-world prognostic utility, the main evaluation of the predictive analyses focused on the subset of patients who did not present motor complications at baseline.

### Preprocessing and statistical analyses

As initial preprocessing step, variables with a missing data proportion exceeding 50% were discarded. The remaining features then underwent a baseline statistical comparison between the clinical outcome groups (LID and motor fluctuations). This comparison, presented in Table S1, was performed using a complete-case analysis, applying the Mann-Whitney U test for continuous data and Pearson’s chi-square test for categorical data. Before each training session, the remaining missing values were imputed using the median for numerical features and the mode (most frequent category) for categorical features; then, all numerical features were scaled to a mean of 0 and a standard deviation of 1.

### Training

Binary classification of patients was performed to predict the onset of the target outcome (LID or motor fluctuations) in PD patients within three years from baseline evaluations. Five ML models were tested: Random Forest (RF), ExtraTrees Classifier (ETC), Extreme Gradient Boosting (XGB), Logistic Regression (LR), Support Vector Classifier (SVC). In addition, two ensemble techniques were employed: a Voting classifier that aggregated predictions from RF, SVC, XGB, and LR, and a Stacking classifier that used the predictions from XGB, RF, and SVC as input features for a final LR meta-model.

To assess model performance, a Randomized Nested Grid Search Cross Validation (CV) strategy was implemented; the workflow is summarized in Figure 3. The full procedure iterated 30 times to assess stability, with the average and standard deviations across these iterations reported as results. In each of the 30 iterations, the entire dataset was first randomly split into 80% for training and 20% for hold-out testing. To ensure representative data splits, this partitioning was stratified by both the target outcome and the baseline symptom status (i.e., ensuring an equal proportion of patients with baseline motor complications in both the training and test sets).

**Figure 3.**
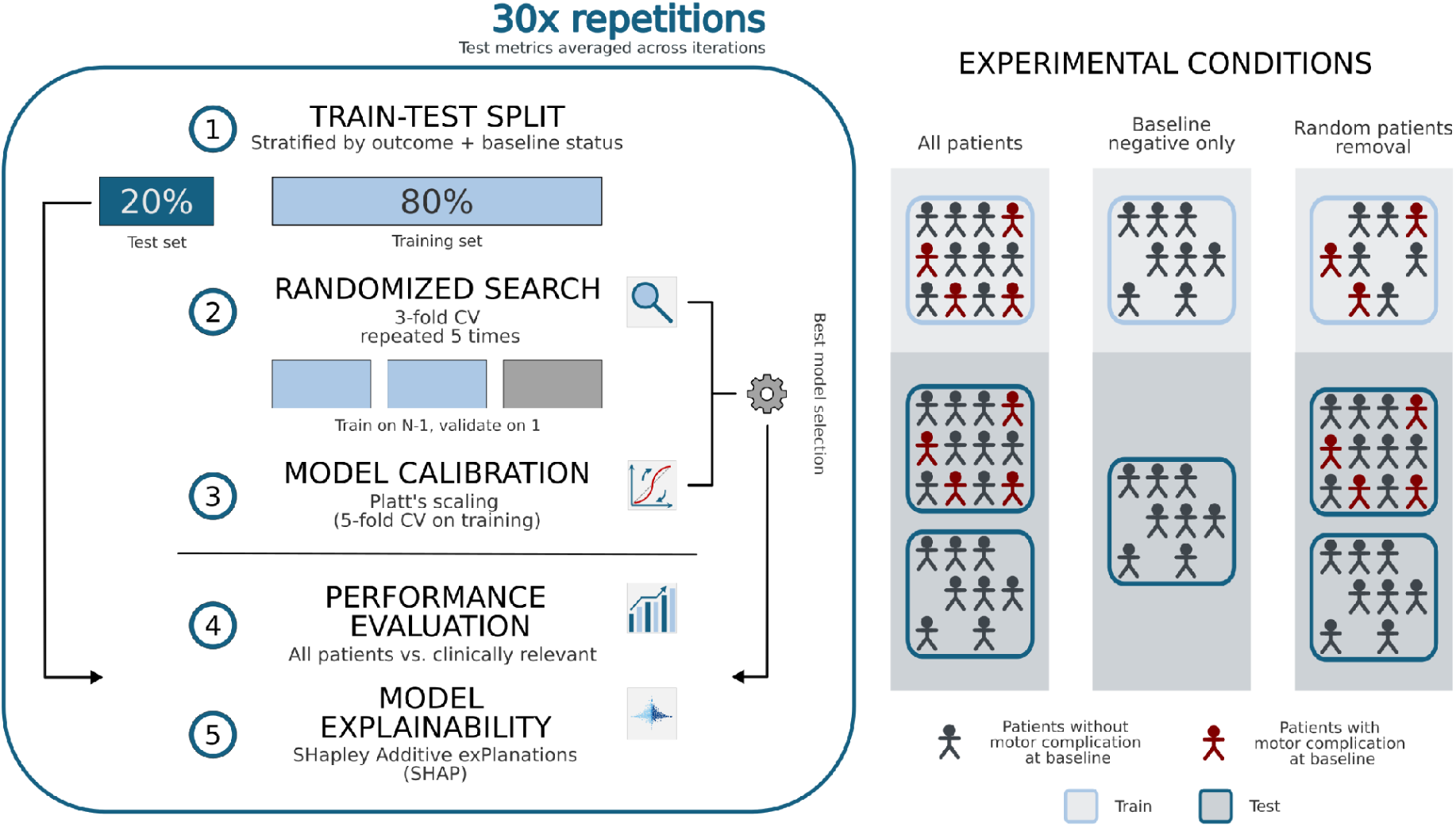
Schematic representation of the repeated nested cross-validation pipeline and experimental conditions. The entire workflow was repeated 30 times, reporting mean ± SD across iterations. Left panel. (1) Train–test split: in each outer iteration, data were split into 80% training and 20% hold-out test, stratified by outcome and baseline motor-complication status. (2) Randomized search: within the training set, hyperparameters were optimized via a randomized search (20 configurations; Table S2) using 3-fold CV (5 repeats), selecting the configuration with the best mean MCC across internal validation folds. (3) Model calibration: the selected model was probability-calibrated with Platt scaling using 5-fold CV on the training data (see Supplementary Materials, Figures S1, S3). (4) Performance evaluation: metrics (MCC, AUC-ROC, F1-score, sensitivity, specificity) were computed on the hold-out test set both globally (all patients) and in the clinically relevant subgroup excluding patients with baseline LID/motor fluctuations. (5) Model explainability: calibrated risk predictions were interpreted with SHAP, aggregating explanations across outer iterations to assess stability of feature contributions. Right panel. Experimental conditions used to probe the role of baseline motor-complication status in learning: training on all patients, training on baseline-negative only, and a random patient-removal control (matched in sample size and preserving the stratification of both outcome and baseline motor complications). CV: cross validation; MCC: Matthew’s Correlation Coefficient; LID: Levodopa induced dyskinesia.

Within each of the 30 training loops, a repeated (5 repetitions) 3-fold CV approach was employed to find the best model configuration over a predefined parameter space (randomized grid search over 20 parameters combinations; see Table S2). The model configuration achieving the best averaged Matthews Correlation Coefficient (MCC) (15) across these internal validation folds was selected as the optimal architecture for that iteration and refitted on the entire training set.

To obtain better-aligned probability estimates, the optimal model underwent probability calibration prior to final evaluation. Raw classifier outputs were mapped to true posterior probabilities using an internal CV strategy. Specifically, Platt Scaling (16) was applied via a 5-fold CV scheme on the training data. Calibration quality was assessed according to the calibration hierarchy proposed by Van Calster et al. (17): *weak calibration* was quantified via the calibration intercept and slope, while *moderate calibration* was visually inspected using flexible calibration curves to ensure alignment between predicted probabilities and observed frequencies (see Supplementary Materials, Figure S1 and S3).

All model training and optimization procedures were implemented using scikit-learn. All computational steps, including preprocessing and classification, were integrated into a single pipeline to create a clean and reproducible training flow and to prevent data leakage (i.e., ensure that information from the test set did not influence the training process).

### Evaluation

Model performance was assessed with a strong emphasis on clinical utility. As described in the training settings, ML classifiers were developed on the entire training population to maximize available information utilization. Performance metrics (MCC, AUC-ROC, F1-Score, Sensitivity and Specificity) were computed on the hold-out test set in two distinct ways:

● Global Evaluation: metrics were calculated on all patients in the test set. These results were inherently biased due to the inclusion of patients who already had the target motor complication at baseline.
● Clinically Relevant Evaluation: all patients presenting LID or motor fluctuations at baseline were excluded from the computation of performance metrics. This analysis specifically assesses the model’s ability to forecast the onset of motor complication in patients that did not present LID or motor fluctuations at baseline - the clinically relevant scenario.

Two key methodological decisions underpinned this evaluation framework. First, to ensure the models were evaluated on their ability to predict the onset of LID or motor fluctuations, the baseline value of the target outcome was deliberately excluded from the feature set. This prevented the models from learning a trivial classification shortcut based on the presence or absence of the target outcome at baseline.

Second, to further examine the contribution of baseline motor complications to model learning, two complementary analyses were performed: retraining the models on the subset of patients without LID or motor fluctuations at baseline, and a controlled ablation experiment designed to separate the effect of excluding these patients from that of reduced sample size. Full methodological details are provided in Methods, “Ablation study” subsection.

### Model explainability

To ensure clinical transparency, model explainability was assessed using SHAP (SHapley Additive exPlanations) (18) to interpret the calibrated risk predictions. SHAP values were computed directly on the final probability outputs using a model-agnostic Kernel Explainer ensuring that feature contributions reflected the actual clinical risk scores. This analysis was performed iteratively across each outer iteration, generating local explanations for the held-out test set. These values were subsequently concatenated to produce a single global summary plot, visualizing the stability and magnitude of feature contributions across the entire study cohort.

Beyond the global feature rankings, the contribution of LEDD to the prediction of future LID was specifically investigated. The aim was to determine how variations in feature value influenced this prediction, and to identify a critical dosage threshold. To explore this non-linear relationship, SHAP dependence plots were generated to illustrate the marginal contribution (SHAP value) of LEDD across its observed values in the patient cohort.

### Ablation study: assessing the role of baseline motor complications

To investigate the specific contribution of patients presenting with Levodopa-induced dyskinesia (LID) or motor fluctuations at baseline to the models’ overall learning process, two complementary analyses were performed. Both procedures strictly adhered to the Nested Grid Search CV pipeline previously described.

First, the models were retrained solely on the subset of patients who were free from LID or motor fluctuations at baseline. This analysis aimed to determine whether the model could effectively learn onset trajectories by relying exclusively on the feature signals of patients without pre-existing motor complications.

Because excluding patients with baseline complications inherently reduces the training set size, an ablation experiment was designed to disentangle the effect of sample-size reduction (quantity of data) from the effect of losing informational signals from patients with baseline motor complications (quality of signal). In each outer iteration of the cross-validation, a random subset of patients from the full training set was removed. The size of this randomly removed group was kept strictly equal to the size of the excluded LID or motor fluctuation group. Crucially, this random removal preserved the original stratification of both the target outcomes and the baseline motor complications.

By comparing performance across the full training set, the restricted incident-only subset, and the randomly reduced control set, it was possible to determine whether performance variations were primarily driven by fewer training examples or by the specific loss of informative signals from patients with baseline complications.

## Data availability

The data that support the findings of this study are available on request from the corresponding author. The data are not publicly available due to privacy or ethical restrictions.

## Code availability

The code used to perform the analyses and generate the results reported in this study is publicly available at https://github.com/endrwalter/pd_motor_symptoms.

## Acknowledgments

The authors thank all the patients involved in the NeuroArtP3 project (NET-2018-12366666), as well as colleagues from the different institutions and departments that contributed to this initiative: Andrea Falini, Antonella Castellano, Andrea Rossi, Domenico Tortora, Costanza Parodi, Antonio Verrico, Federica Sabatini, Emilio Portaccio, Matteo Betti, Guido Pasquini, Filippo Gerli, Claudia Niccolai, Isabella Cama, Cristina Campi, Alessio Cirone, Sara Garbarino, Michele Piana, Donatella Ottaviani, Raffaella Di Giacopo, Ruggero Bacchin.

This research was funded by the Italian Ministry of Health, grant number NET-2018-12366666 (NeuroArtP3). This research was partially supported by #NEXTGENERATIONEU (NGEU) and funded by the Ministry of University and Research (MUR), National Recovery and Resilience Plan (NRRP), project MNESYS (PE0000006)—(DN. 1553 11.10.2022).

## Author contributions

W.E.: Data curation, Formal analysis, Writing – original draft, Writing – review & editing. N.C.: Investigation, Writing – original draft, Writing – review & editing. C. L.: Conceptualization, Data curation, Investigation, Writing – original draft, Writing – review & editing. M.M.: Data curation, Formal analysis, Writing – original draft, Writing – review & editing. F.R.: Data curation, Formal analysis, Writing – original draft, Writing – review & editing. S.B.: Data curation, Formal analysis, Writing – original draft, Writing – review & editing. L. G.: Project administration, Writing – review & editing. A.U.: Conceptualization, Project administration, Writing – review & editing. B. G.: Conceptualization, Writing – review & editing. G. J.: Conceptualization, Formal analysis, Supervision, Writing – review & editing. V.O.: Conceptualization, Formal analysis, Writing – original draft, Writing – review & editing. M. C.M.: Conceptualization, Investigation, Methodology, Validation, Writing – review & editing.

## Author Information

NeuroArtP3 Network is composed by: Andrea Falini^11,12^, Antonella Castellano^11,12^, Andrea Rossi^13,14^, Domenico Tortora^13^, Costanza Parodi^13^, Antonio Verrico^15^, Federica Sabatini^16^, Emilio Portaccio^17^, Matteo Betti^17^, Guido Pasquini^18^, Filippo Gerli^18^, Claudia Niccolai^18^, Isabella Cama^19,20^, Cristina Campi^19,20^, Alessio Cirone^19,21^, Sara Garbarino^19,21^, Michele Piana^19,20^, Donatella Ottaviani^22^, Raffaella Di Giacopo^22^, Ruggero Bacchin^22^

^11^Neuroradiology Unit and CERMAC, IRCCS Ospedale San Raffaele, Milan, Italy

^12^Università Vita-Salute San Raffaele, Milan, Italy

^13^Neuroradiology Unit, IRCCS Istituto Giannina Gaslini, Genoa, Italy

^14^Department of Health Sciences (DISSAL), University of Genoa, Genoa, Italy

^15^Neuro-Oncology Unit, IRCCS Istituto Giannina Gaslini, Genoa, Italy

^16^UOSD Cell Factory, IRCCS Istituto Giannina Gaslini, Genoa, Italy

^17^Department of Neuroscience, Psychology, Drug Research and Child Health (NEUROFARBA), University of Florence, Florence, Italy

^18^IRCCS Don Carlo Gnocchi Foundation, Florence, Italy

^19^Life Science Computational Laboratory (LISCOMP), IRCCS Ospedale Policlinico San Martino, Genoa, Italy

^20^Department of Mathematics (DIMA), University of Genoa, Genoa, Italy

^21^IRCCS Ospedale Policlinico San Martino, Genoa, Italy

^22^Azienda sanitaria universitaria integrata del Trentino (ASUIT), Trento, Italy

## References

1. Freitas ME, Hess CW, Fox SH. Motor complications of dopaminergic medications in Parkinson’s disease. Semin Neurol. 2017 Apr;37(2):147–57.

2. Kim HJ, Mason S, Foltynie T, Winder-Rhodes S, Barker RA, Williams-Gray CH. Motor complications in Parkinson’s disease: 13-year follow-up of the CamPaIGN cohort. Mov Disord. 2020 Jan;35(1):185–90.

3. Anderson E, Nutt J. The long-duration response to levodopa: phenomenology, potential mechanisms and clinical implications. Parkinsonism Relat Disord. 2011 Sept 1;17(8):587–92.

4. Cilia R, Akpalu A, Sarfo FS, Cham M, Amboni M, Cereda E, et al. The modern pre-levodopa era of Parkinson’s disease: insights into motor complications from sub-Saharan Africa. Brain. 2014 Oct;137(Pt 10):2731–42.

5. Huang Q, Weng H, Weng X, Zhao ZH. Selected risk factors for motor fluctuations in Parkinson’s disease: a meta-analysis. Clin Neurol Neurosurg. 2025 Aug;255(108921):108921.

6. Sharma JC, Bachmann CG, Linazasoro G. Classifying risk factors for dyskinesia in Parkinson’s disease. Parkinsonism Relat Disord. 2010 Sept;16(8):490–7.

7. Péchevis M, Clarke CE, Vieregge P, Khoshnood B, Deschaseaux-Voinet C, Berdeaux G, et al. Effects of dyskinesias in Parkinson’s disease on quality of life and health-related costs: a prospective European study. Eur J Neurol. 2005 Dec 1;12(12):956–63.

8. Aradi SD, Hauser RA. Medical management and prevention of motor complications in Parkinson’s disease. Neurotherapeutics. 2020 Oct;17(4):1339–65.

9. Loo RTJ, Tsurkalenko O, Klucken J, Mangone G, Khoury F, Vidailhet M, et al. Levodopa-induced dyskinesia in Parkinson’s disease: Insights from cross-cohort prognostic analysis using machine learning. Parkinsonism Relat Disord. 2024 Sept 1;126(107054):107054.

10. Martorell-Marugán J, Carmona-Sáez P, Chierici M, Bandres-Ciga S, Jurman G. Machine Learning applications in the study of Parkinson’s disease: A systematic review. Curr Bioinform. 2023 Aug 1;18(7):576–86.

11. Loo RTJ, Pavelka L, Mangone G, Khoury F, Vidailhet M, Corvol JC, et al. Interpretable machine learning for cross-cohort prediction of motor fluctuations in Parkinson’s disease. Mov Disord. 2025 Aug;40(8):1604–17.

12. Malaguti MC, Gios L, Giometto B, Longo C, Riello M, Ottaviani D, et al. Artificial intelligence of imaging and clinical neurological data for predictive, preventive and personalized (P3) medicine for Parkinson Disease: The NeuroArtP3 protocol for a multi-center research study. PLoS One. 2024 Mar 14;19(3):e0300127.

13. Postuma RB, Berg D, Stern M, Poewe W, Olanow CW, Oertel W, et al. MDS clinical diagnostic criteria for Parkinson’s disease: MDS-PD Clinical Diagnostic Criteria. Mov Disord. 2015 Oct;30(12):1591–601.

14. Jankovic J. Motor fluctuations and dyskinesias in Parkinson’s disease: clinical manifestations. Mov Disord. 2005;20 Suppl 11(S11):S11–6.

15. Chicco D, Jurman G. The Matthews correlation coefficient (MCC) should replace the ROC AUC as the standard metric for assessing binary classification. BioData Min. 2023 Feb 17;16(1):4.

16. Platt J. Probabilistic Outputs for Support Vector Machines and Comparisons to Regularized Likelihood Methods. In: Advances in Large Margin Classifiers. London, England: MIT Press; 1999.

17. Van Calster B, McLernon DJ, van Smeden M, Wynants L, Steyerberg EW, Topic Group ‘Evaluating diagnostic tests and prediction models’ of the STRATOS initiative. Calibration: the Achilles heel of predictive analytics. BMC Med. 2019 Dec 16;17(1):230.

18. Lundberg S, Lee SI. A unified approach to interpreting model predictions [Internet]. arXiv [cs.AI]. 2017 [cited 2026 Feb 27]. Available from: 10.5555/3295222.3295230

19. Warren Olanow C, Kieburtz K, Rascol O, Poewe W, Schapira AH, Emre M, et al. Factors predictive of the development of Levodopa-induced dyskinesia and wearing-off in Parkinson’s disease: Risk Factors forL-Dopa Induced Motor Complications. Mov Disord. 2013 July 1;28(8):1064–71.

20. Eusebi P, Romoli M, Paoletti FP, Tambasco N, Calabresi P, Parnetti L. Risk factors of levodopa-induced dyskinesia in Parkinson’s disease: results from the PPMI cohort. NPJ Parkinsons Dis. 2018 Nov 16;4(1):33.

21. Scott NW, Macleod AD, Counsell CE. Motor complications in an incident Parkinson’s disease cohort. Eur J Neurol. 2016 Feb 1;23(2):304–12.

22. Conti V, Izzo V, Russillo MC, Picillo M, Amboni M, Scaglione CLM, et al. Gender differences in levodopa pharmacokinetics in levodopa-naïve patients with Parkinson’s disease. Front Med (Lausanne). 2022 May 31;9:909936.

23. Schrag A, Quinn N. Dyskinesias and motor fluctuations in Parkinson’s disease. A community-based study. Brain. 2000 Nov;123 ( Pt 11):2297–305.

24. Kostic V, Przedborski S, Flaster E, Sternic N. Early development of levodopa-induced dyskinesias and response fluctuations in young-onset Parkinson’s disease. Neurology. 1991 Feb 1;41(2 ( Pt 1)):202–5.

25. Grandas F, Galiano ML, Tabernero C. Risk factors for levodopa-induced dyskinesias in Parkinson’s disease. J Neurol. 1999 Dec;246(12):1127–33.

26. de la Fuente-Fernández R, Sossi V, Huang Z, Furtado S, Lu JQ, Calne DB, et al. Levodopa-induced changes in synaptic dopamine levels increase with progression of Parkinson’s disease: implications for dyskinesias. Brain. 2004 Dec 25;127(Pt 12):2747–54.

27. Chung SJ, Lee HS, Yoo HS, Lee YH, Lee PH, Sohn YH. Patterns of striatal dopamine depletion in early Parkinson disease: Prognostic relevance: Prognostic relevance. Neurology. 2020 July 21;95(3):e280–90.

28. Bjornestad A, Forsaa EB, Pedersen KF, Tysnes OB, Larsen JP, Alves G. Risk and course of motor complications in a population-based incident Parkinson’s disease cohort. Parkinsonism Relat Disord. 2016 Jan 1;22:48–53.

29. Hung AY, Schwarzschild MA. Treatment of Parkinson’s disease: what’s in the non-dopaminergic pipeline? Neurotherapeutics. 2014 Jan;11(1):34–46.

30. Aslam S, Manfredsson F, Stokes A, Shill H. “Advanced” Parkinson’s disease: A review. Parkinsonism Relat Disord. 2024 June;123(106065):106065.

31. de Bie RMA, Katzenschlager R, Swinnen BEKS, Peball M, Lim SY, Mestre TA, et al. Update on treatments for Parkinson’s disease motor fluctuations - an International Parkinson and Movement Disorder Society Evidence-Based Medicine review. Mov Disord. 2025 May;40(5):776–94.

