## Supplementary Materials for "Explainable machine learning reveals the challenges of predicting new-onset motor complications in Parkinson’s disease"

| | Future LID | | | Future motor fluctuations | | |
|---|---|---|---|---|---|---|
| **Attribute** | **Negative** | **Positive** | **P-value** | **Negative** | **Positive** | **P-value** |
| Use of DA (No) N (%) | 48 (31.8%) | 26 (27.1%) | 0.519 | 41 (30.1%) | 33 (29.2%) | 0.921 |
| Anxiety (Absent) N (%) | 119 (78.8%) | 67 (69.8%) | 0.147 | 102 (76.1%) | 84 (74.3%) | 0.861 |
| Cognitive status (Normal) N (%) | 134 (88.7%) | 83 (86.6%) | 0.635 | 119 (88.8%) | 98 (86.7%) | 0.938 |
| Cognitive status (MCI) N (%) | 10 (6.6%) | 9 (9.4%) | | 11 (8.2%) | 8 (7.1%) | |
| Cognitive status (Dementia) N (%) | 3 (2.0%) | 3 (3.1%) | | 3 (2.2%) | 3 (2.7%) | |
| Depression (Absent) N (%) | 107 (70.9%) | 70 (72.9%) | 0.838 | 84 (62.7%) | 93 (82.3%) | 0.001 |
| Presence of dysautonomia (Absent) N (%) | 92 (60.9%) | 67 (69.8%) | 0.166 | 82 (61.2%) | 77 (68.1%) | 0.006 |
| Presence of dysautonomia (Other) N (%) | 48 (31.8%) | 20 (20.8%) | | 46 (34.3%) | 22 (19.5%) | |
| Presence of dysautonomia (Orthostatic hypotension) N (%) | 11 (7.3%) | 9 (9.4%) | | 6 (4.5%) | 14 (12.4%) | |
| Disease duration (0-5) N (%) | 106 (70.2%) | 39 (40.6%) | < 0.001 | 96 (71.6%) | 49 (43.4%) | < 0.001 |
| Disease duration (5-10) N (%) | 34 (22.5%) | 29 (30.2%) | | 29 (21.6%) | 34 (30.1%) | |
| Disease duration (>10) N (%) | 11 (7.3%) | 28 (29.2%) | | 9 (6.7%) | 30 (26.5%) | |
| LID (Absent) N (%) | 151 (100.0%) | 62 (64.6%) | < 0.001 | 132 (97.8%) | 82 (72.6%) | < 0.001 |
| Falls (Absent) N (%) | 136 (90.1%) | 80 (83.3%) | 0.174 | 120 (89.6%) | 96 (85.0%) | 0.372 |
| Family history (for PD) (No) N (%) | 101 (66.2%) | 67 (69.8%) | 0.657 | 83 (61.9%) | 84 (74.3%) | 0.053 |
| Freezing (Absent) N (%) | 137 (90.7%) | 79 (82.3%) | 0.079 | 123 (91.8%) | 93 (82.3%) | 0.040 |
| Gender (Male) N (%) | 100 (66.2%) | 51 (53.1%) | 0.054 | 86 (64.2%) | 65 (57.5%) | 0.348 |
| H&Y stage 2 N (%) | 76 (50.3%) | 52 (54.2%) | 0.179 | 70 (52.2%) | 58 (51.3%) | 0.098 |
| H&Y stage 1 N (%) | 53 (35.1%) | 24 (25.0%) | | 47 (35.1%) | 30 (26.5%) | |
| H&Y stage >2 N (%) | 22 (14.6%) | 20 (20.8%) | | 17 (12.7%) | 25 (22.1%) | |
| Hyposmia N (%) | 73 (48.3%) | 64 (66.7%) | 0.005 | 64 (47.8%) | 73 (64.6%) | 0.006 |
| ICD N (%) | 142 (94.0%) | 85 (88.5%) | 0.192 | 125 (93.3%) | 102 (90.3%) | 0.527 |
| Use of MAOIs (N) (%) | 59 (39.1%) | 41 (42.7%) | 0.664 | 43 (32.1%) | 57 (50.4%) | 0.005 |
| Use of Levodopa N (%) | 61 (40.4%) | 14 (14.6%) | < 0.001 | 61 (45.5%) | 14 (12.4%) | < 0.001 |
| Motor fluctuations (Absent) N (%) | 139 (92.1%) | 49 (51.0%) | < 0.001 | 134 (100.0%) | 54 (47.8%) | < 0.001 |

| | | | | | | |
|---|---|---|---|---|---|---|
| Phenotype (Akinetic) N (%) | 56 (37.1%) | 46 (47.9%) | 0.121 | 51 (38.1%) | 51 (45.1%) | 0.320 |
| Sleep/Wake disorders (Absent) N (%) | 84 (55.6%) | 62 (64.6%) | 0.207 | 73 (54.5%) | 73 (64.6%) | 0.138 |
| Age at baseline Median [Q1, Q3] | 70 [63, 75] | 68 [61.75, 73] | 0.061 | 70 [64.25, 75] | 67 [61, 73] | 0.024 |
| DA use, duration Median [Q1, Q3] | 0 [0, 3] | 3 [0, 7] | < 0.001 | 0 [0, 3] | 3 [0, 7] | < 0.001 |
| Education Median [Q1, Q3] | 11.5 [8, 13] | 9 [8, 13] | 0.407 | 13 [8, 13] | 10 [8, 13] | 0.212 |
| IMAOYears Median [Q1, Q3] | 0 [0, 1] | 0 [0, 2] | 0.020 | 0 [0, 1] | 0 [0, 2] | 0.174 |
| Levodopa use, duration Median [Q1, Q3] | 0 [0, 2] | 4 [2, 8] | < 0.001 | 0 [0, 2] | 3.5 [2, 7] | < 0.001 |
| MDS-UPDRS III (ON) Median [Q1, Q3] | 19 [11, 29] | 16 [12, 21.75] | 0.081 | 19 [12.5, 29] | 16 [11, 22] | 0.032 |
| Time between symptoms onset and diagnosis Median [Q1, Q3] | 1 [0, 2] | 1 [0, 2] | 0.952 | 1 [0, 2] | 1 [0, 2] | 0.775 |
| LEDD Median [Q1, Q3] | 200 [0, 300] | 400 [275, 650] | < 0.001 | 150 [0, 300] | 400 [250, 625] | < 0.001 |

**Table S1: Descriptive statistics of baseline attributes and statistical significance with respect to the outcome.** Baseline characteristics stratified by future outcome. Continuous variables are presented as median [Q1, Q3] and were compared using the Mann-Whitney U test. Categorical variables are presented as count (percentage of group total) and were compared using Pearson's chi-square test. For multi-level categorical variables, the p-value tests the overall distribution. P-values <0.001 indicate strong statistical evidence. Statistical tests were performed using complete-case analysis, excluding missing values. DA: dopamine agonist; H&Y: Hoehn and Yahr stage; ICD: impulse control disorder; LEDD: levodopa equivalent daily dose; LID: Levodopa-induced dyskinesia; MAOIs: monoamine oxidase inhibitors.

| Model | Hyperparameter | Search Space / Values |
|---|---|---|
| Random Forest | max_depth | 1 to 110 (step 1) |
| | min_samples_split | 2 to 30 (step 1) |
| | max_features | log2, sqrt |
| | n_estimators | 100 to 1000 (step 5) |
| | class_weight | balanced, balanced_subsample |
| Extra Trees | n_estimators | 1 to 800 (step 5) |
| | max_depth | 1 to 40 (step 1) |
| | max_features | 1 to 40 (step 1) |
| | class_weight | balanced, balanced_subsample |
| XGBoost | learning_rate | 0.01 to 0.1 (Uniform) |
| | subsample | 0.4 to 1.0 (Uniform) |
| | n_estimators | 50 to 250 (Integer) |
| | max_depth | 1 to 20 (step 1) |
| | gamma | 0 to 0.5 (Uniform) |
| | scale_pos_weight | 1, 2, 3, 4, 5, 6, 7, 8, 9 |
| Logistic Regression | C | 0.001 to 1000 (Log-Uniform) |
| | max_iter | 500 to 1000 (step 1) |
| | fit_intercept | True, False |
| | class_weight | balanced |
| SVC | C | 0.0001 to 10000 (Log-Uniform) |
| | kernel | linear, poly, rbf, sigmoid |
| | degree | 2, 3, 4 |
| | gamma | 0.001 to 1.0 (Uniform) |
| | coef0 | 0, 0.5, 1 |
| | shrinking | True, False |
| | class_weight | balanced |

**Table S2.** Summary of the hyperparameter search spaces and sampling strategies used for model optimization. Ranges for continuous variables are specified with their respective distribution types (Uniform or Log-Uniform), while discrete parameters include the step size or the set of specific categorical values evaluated during the tuning process.

| Train cohort composition | | All | | Non dyskinetic only | All (r. s.) | |
|---|---|---|---|---|---|---|
| Test cohort composition | | All | Non dyskinetic only | Non dyskinetic only | All (r. s.) | Non dyskinetic only (r. s.) |
| Model | Metric | | | | | |
| ETC | AUC | 0.77 ± 0.06 | 0.70 ± 0.08 | 0.67 ± 0.08 | 0.78 ± 0.06 | 0.71 ± 0.08 |
| | F1 | 0.59 ± 0.07 | 0.40 ± 0.10 | 0.11 ± 0.12 | 0.60 ± 0.08 | 0.43 ± 0.16 |
| | MCC | 0.42 ± 0.11 | 0.24 ± 0.14 | 0.03 ± 0.15 | 0.45 ± 0.11 | 0.29 ± 0.18 |
| | Sens. | 0.51 ± 0.08 | 0.33 ± 0.10 | 0.07 ± 0.09 | 0.50 ± 0.10 | 0.35 ± 0.15 |
| | Spec. | 0.87 ± 0.07 | 0.87 ± 0.07 | 0.95 ± 0.05 | 0.90 ± 0.05 | 0.90 ± 0.05 |
| | PPV | 0.72 ± 0.11 | 0.53 ± 0.15 | 0.38 ± 0.37 | 0.77 ± 0.09 | 0.58 ± 0.19 |
| | NPV | 0.74 ± 0.03 | 0.77 ± 0.03 | 0.70 ± 0.02 | 0.74 ± 0.04 | 0.77 ± 0.04 |
| LR | AUC | 0.78 ± 0.06 | 0.70 ± 0.07 | 0.69 ± 0.07 | 0.78 ± 0.06 | 0.71 ± 0.09 |
| | F1 | 0.57 ± 0.08 | 0.39 ± 0.11 | 0.21 ± 0.12 | 0.56 ± 0.11 | 0.40 ± 0.14 |
| | MCC | 0.41 ± 0.11 | 0.25 ± 0.13 | 0.17 ± 0.15 | 0.39 ± 0.12 | 0.25 ± 0.15 |
| | Sens. | 0.48 ± 0.10 | 0.33 ± 0.12 | 0.14 ± 0.08 | 0.47 ± 0.13 | 0.34 ± 0.14 |
| | Spec. | 0.88 ± 0.07 | 0.88 ± 0.07 | 0.96 ± 0.05 | 0.88 ± 0.05 | 0.88 ± 0.05 |
| | PPV | 0.72 ± 0.12 | 0.54 ± 0.16 | 0.65 ± 0.29 | 0.71 ± 0.09 | 0.53 ± 0.13 |
| | NPV | 0.74 ± 0.04 | 0.77 ± 0.03 | 0.72 ± 0.02 | 0.72 ± 0.05 | 0.76 ± 0.04 |
| RF | AUC | 0.80 ± 0.06 | 0.73 ± 0.08 | 0.70 ± 0.07 | 0.80 ± 0.06 | 0.73 ± 0.09 |
| | F1 | 0.62 ± 0.07 | 0.45 ± 0.10 | 0.28 ± 0.14 | 0.66 ± 0.06 | 0.51 ± 0.11 |
| | MCC | 0.43 ± 0.11 | 0.27 ± 0.14 | 0.14 ± 0.16 | 0.47 ± 0.09 | 0.34 ± 0.14 |
| | Sens. | 0.59 ± 0.09 | 0.43 ± 0.11 | 0.22 ± 0.11 | 0.61 ± 0.09 | 0.48 ± 0.14 |
| | Spec. | 0.82 ± 0.08 | 0.82 ± 0.08 | 0.89 ± 0.07 | 0.84 ± 0.07 | 0.84 ± 0.07 |
| | PPV | 0.68 ± 0.10 | 0.50 ± 0.14 | 0.47 ± 0.23 | 0.73 ± 0.09 | 0.58 ± 0.12 |
| | NPV | 0.77 ± 0.04 | ,0.79 ± 0.03 | 0.73 ± 0.03 | 0.77 ± 0.04 | 0.79 ± 0.04 |
| STACKING | AUC | 0.79 ± 0.06 | 0.72 ± 0.08 | 0.69 ± 0.06 | 0.79 ± 0.06 | 0.72 ± 0.09 |
| | F1 | 0.62 ± 0.07 | 0.45 ± 0.10 | 0.23 ± 0.14 | 0.64 ± 0.07 | 0.49 ± 0.09 |

| | | | | | | |
|---|---|---|---|---|---|---|
| | MCC | 0.42 ± 0.12 | 0.27 ± 0.14 | 0.12 ± 0.14 | 0.44 ± 0.10 | 0.31 ± 0.13 |
| | Sens. | 0.59 ± 0.09 | 0.44 ± 0.12 | 0.17 ± 0.12 | 0.59 ± 0.09 | 0.45 ± 0.12 |
| | Spec. | 0.82 ± 0.08 | 0.82 ± 0.08 | 0.92 ± 0.06 | 0.83 ± 0.06 | 0.83 ± 0.06 |
| | PPV | 0.67 ± 0.11 | 0.50 ± 0.15 | 0.47 ± 0.19 | 0.71 ± 0.08 | 0.55 ± 0.12 |
| | NPV | 0.77 ± 0.04 | 0.79 ± 0.04 | 0.72 ± 0.03 | 0.76 ± 0.04 | 0.78 ± 0.04 |
| SVC | AUC | 0.79 ± 0.06 | 0.70 ± 0.08 | 0.67 ± 0.08 | 0.78 ± 0.06 | 0.70 ± 0.09 |
| | F1 | 0.61 ± 0.07 | 0.43 ± 0.11 | 0.19 ± 0.14 | 0.60 ± 0.08 | 0.42 ± 0.11 |
| | MCC | **0.44 ± 0.12** | **0.28 ± 0.14** | **0.14 ± 0.17** | **0.43 ± 0.10** | **0.26 ± 0.14** |
| | Sens. | **0.54 ± 0.09** | **0.37 ± 0.12** | **0.12 ± 0.10** | **0.52 ± 0.09** | **0.35 ± 0.11** |
| | Spec. | 0.87 ± 0.07 | 0.87 ± 0.07 | 0.95 ± 0.05 | 0.88 ± 0.06 | 0.88 ± 0.06 |
| | PPV | 0.73 ± 0.12 | 0.55 ± 0.16 | 0.59 ± 0.36 | 0.74 ± 0.09 | 0.56 ± 0.15 |
| | NPV | 0.75 ± 0.04 | 0.78 ± 0.03 | 0.72 ± 0.02 | 0.74 ± 0.04 | 0.76 ± 0.03 |
| VOTING | AUC | 0.80 ± 0.06 | 0.72 ± 0.08 | 0.68 ± 0.08 | 0.79 ± 0.06 | 0.72 ± 0.09 |
| | F1 | 0.63 ± 0.06 | 0.45 ± 0.10 | 0.25 ± 0.14 | 0.64 ± 0.07 | 0.48 ± 0.12 |
| | MCC | 0.43 ± 0.10 | 0.27 ± 0.13 | 0.13 ± 0.16 | 0.45 ± 0.10 | 0.31 ± 0.15 |
| | Sens. | 0.59 ± 0.09 | 0.43 ± 0.12 | 0.18 ± 0.11 | 0.59 ± 0.09 | 0.45 ± 0.14 |
| | Spec. | 0.82 ± 0.08 | 0.82 ± 0.08 | 0.91 ± 0.07 | 0.84 ± 0.06 | 0.84 ± 0.06 |
| | PPV | 0.68 ± 0.10 | 0.50 ± 0.14 | 0.50 ± 0.25 | 0.72 ± 0.09 | 0.55 ± 0.13 |
| | NPV | 0.77 ± 0.04 | 0.79 ± 0.04 | 0.72 ± 0.03 | 0.76 ± 0.04 | 0.78 ± 0.05 |
| XGB | AUC | 0.77 ± 0.06 | 0.70 ± 0.08 | 0.66 ± 0.07 | 0.78 ± 0.06 | 0.72 ± 0.09 |
| | F1 | 0.60 ± 0.08 | 0.45 ± 0.11 | 0.13 ± 0.11 | 0.63 ± 0.07 | 0.49 ± 0.12 |
| | MCC | **0.39 ± 0.13** | **0.26 ± 0.16** | **0.02 ± 0.11** | **0.43 ± 0.11** | **0.31 ± 0.15** |
| | Sens. | **0.57 ± 0.09** | **0.44 ± 0.11** | **0.09 ± 0.09** | **0.57 ± 0.10** | **0.45 ± 0.16** |
| | Spec. | 0.81 ± 0.08 | 0.81 ± 0.08 | 0.92 ± 0.07 | 0.84 ± 0.07 | 0.84 ± 0.07 |
| | PPV | 0.65 ± 0.11 | 0.48 ± 0.15 | 0.36 ± 0.26 | 0.70 ± 0.09 | 0.55 ± 0.12 |
| | NPV | 0.75 ± 0.04 | 0.79 ± 0.04 | 0.70 ± 0.01 | 0.75 ± 0.05 | 0.79 ± 0.05 |

**Table S3. Comparative Performance of Machine Learning Models for Predicting LID Onset.** The table displays key performance metrics (AUC, F1-score, MCC, Sensitivity, Specificity) under three distinct experimental conditions: models trained on all patients (first two columns), models directly trained only on patients without LID at baseline (third column) and models trained using a random subset of patients matching in size the cohort of non-dyskinetic patients at baseline (columns four and five). Performance is evaluated on both the entire test set (All) and the clinically relevant subset of patients without LID at baseline (Non dyskinetic only). All values are presented as mean ± standard deviation. Sens.: Sensitivity, Spec.: Specificity, r.s.: random subset.

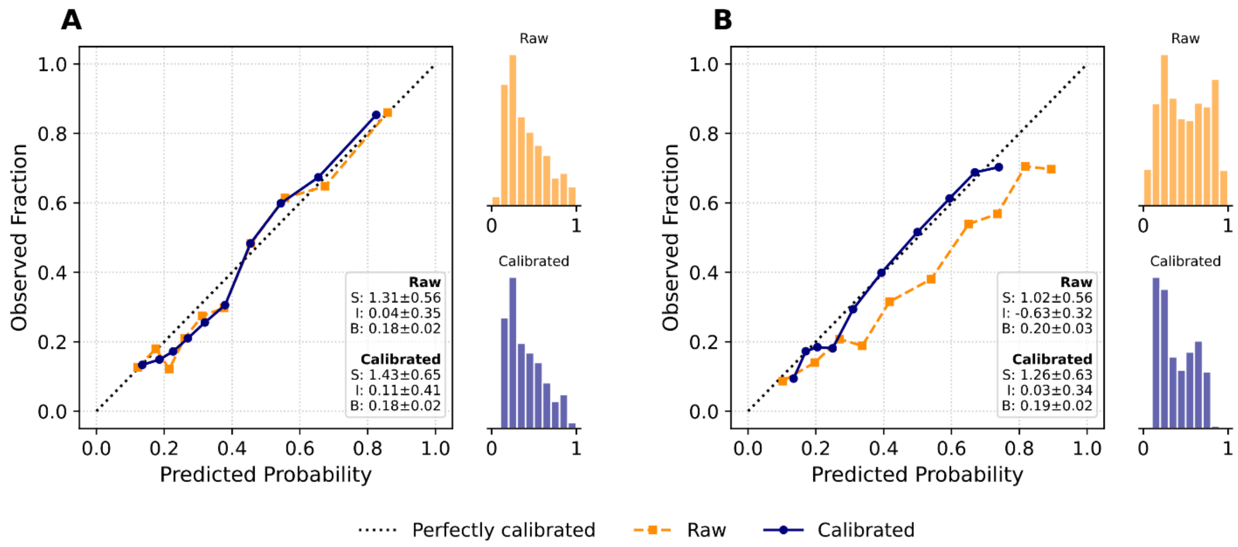


**Figure S1. Probability calibration analysis of the optimal predictive models - LID Task.** The plots illustrate the alignment between predicted probabilities (x-axis) and observed outcome frequencies (y-axis) for the two best-performing models: (A) Support Vector Classifier (SVC) and (B) Extreme Gradient Boosting (XGB). The orange squares (dashed line) represent the raw, uncalibrated classifier outputs, while the dark blue circles (solid line) depict the posterior probabilities after calibration via Platt Scaling. The diagonal dotted line indicates perfect calibration. Quantile binning (equal sample size per bin) was employed to ensure statistical robustness across the probability range. Inset text reports the quantitative calibration metrics: Slope (S, ideal=1), Intercept (I, ideal=0), and Brier Score (B, lower indicates better accuracy). Marginal histograms display the distribution of predicted probabilities, highlighting the shift from raw scores (orange) to calibrated probabilities (blue). Note that the SVC (A) demonstrates naturally higher resolution (predictions clustered at extremes) compared to the XGB model (B).

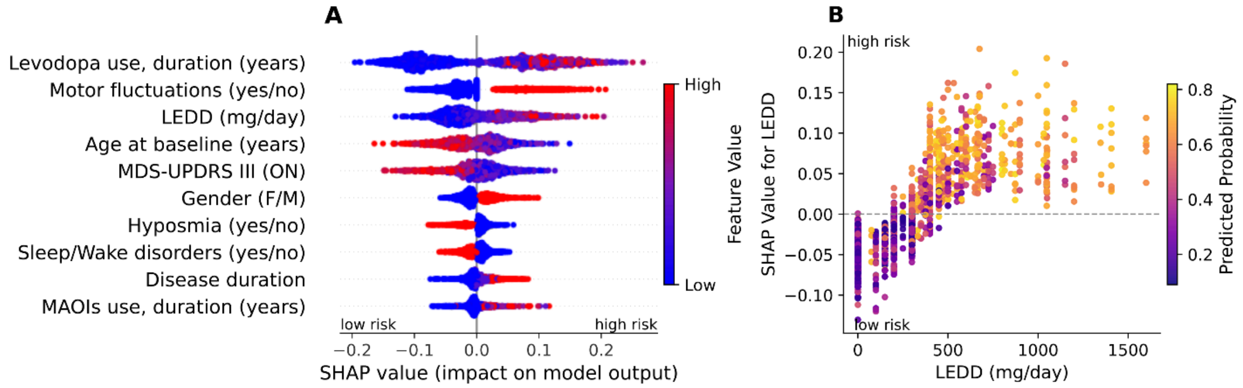


**Figure S2. SHAP analysis of XGB for the prediction of future LID. A.** Global feature importance and impact for the XGB model revealed by SHAP. Features are ranked in descending order of importance. Each dot represents a patient from the test set. The horizontal axis shows the SHAP value, indicating the feature's contribution to predicting LID (positive values) or absence of LID (negative values) within the third year. Color indicates the feature's original value (blue=low, red=high), revealing the direction and magnitude of its effect. **B.** Detailed analysis of the levodopa equivalent daily dose (LEDD) feature using a SHAP dependence plot for the XGB model. The x-axis plots the patient's actual LEDD on its original scale, while the y-axis plots the SHAP value, indicating the feature's contribution to the prediction. A potential dosage threshold is visible around 300-400 mg, where the feature's impact (SHAP value) transitions from negative (low risk) to positive (high risk). LEDD: levodopa equivalent daily dose; MAOIs: monoamine oxidase inhibitors.

| Train cohort composition | | All | | Non fluctuating only | All (r.s.) | |
|---|---|---|---|---|---|---|
| Test cohort composition | | All | Non fluctuating only | Non fluctuating only | All (r.s.) | Non fluctuating only (r.s.) |
| Model | Metric | | | | | |
| ETC | AUC | 0.80 ± 0.06 | 0.72 ± 0.08 | 0.68 ± 0.09 | 0.81 ± 0.06 | 0.72 ± 0.08 |
| | F1 | 0.70 ± 0.06 | 0.49 ± 0.12 | 0.04 ± 0.11 | 0.70 ± 0.06 | 0.49 ± 0.09 |
| | MCC | 0.45 ± 0.10 | 0.28 ± 0.16 | -0.01 ± 0.11 | 0.48 ± 0.11 | 0.31 ± 0.14 |
| | Sens. | 0.69 ± 0.09 | 0.52 ± 0.15 | 0.03 ± 0.08 | 0.70 ± 0.09 | 0.53 ± 0.16 |
| | Spec. | 0.76 ± 0.07 | 0.76 ± 0.07 | 0.97 ± 0.06 | 0.77 ± 0.10 | 0.77 ± 0.10 |
| | PPV | 0.71 ± 0.06 | 0.47 ± 0.11 | 0.21 ± 0.29 | 0.72 ± 0.10 | 0.51 ± 0.16 |
| | NPV | 0.74 ± 0.05 | 0.80 ± 0.05 | 0.71 ± 0.02 | 0.76 ± 0.05 | 0.82 ± 0.05 |
| LR | AUC | 0.82 ± 0.05 | 0.72 ± 0.09 | 0.70 ± 0.09 | 0.83 ± 0.06 | 0.74 ± 0.09 |
| | F1 | 0.71 ± 0.05 | 0.49 ± 0.12 | 0.24 ± 0.13 | 0.70 ± 0.06 | 0.48 ± 0.10 |
| | MCC | 0.51 ± 0.08 | 0.30 ± 0.16 | 0.16 ± 0.17 | 0.50 ± 0.10 | 0.30 ± 0.14 |
| | Sens. | 0.67 ± 0.08 | 0.47 ± 0.14 | 0.16 ± 0.09 | 0.66 ± 0.08 | 0.46 ± 0.12 |
| | Spec. | 0.82 ± 0.06 | 0.82 ± 0.06 | 0.94 ± 0.06 | 0.82 ± 0.08 | 0.82 ± 0.08 |
| | PPV | 0.77 ± 0.06 | 0.52 ± 0.11 | 0.56 ± 0.28 | 0.76 ± 0.09 | 0.53 ± 0.16 |
| | NPV | 0.75 ± 0.04 | 0.79 ± 0.05 | 0.73 ± 0.02 | 0.75 ± 0.04 | 0.80 ± 0.04 |
| RF | AUC | 0.81 ± 0.06 | 0.72 ± 0.09 | 0.72 ± 0.09 | 0.83 ± 0.07 | 0.74 ± 0.10 |
| | F1 | 0.74 ± 0.06 | 0.52 ± 0.13 | 0.18 ± 0.12 | 0.74 ± 0.06 | 0.53 ± 0.11 |
| | MCC | 0.53 ± 0.11 | 0.32 ± 0.18 | 0.10 ± 0.15 | 0.55 ± 0.12 | 0.35 ± 0.15 |
| | Sens. | 0.73 ± 0.08 | 0.53 ± 0.15 | 0.12 ± 0.09 | 0.74 ± 0.08 | 0.55 ± 0.15 |
| | Spec. | 0.79 ± 0.07 | 0.79 ± 0.07 | 0.93 ± 0.06 | 0.80 ± 0.10 | 0.80 ± 0.10 |
| | PPV | 0.75 ± 0.07 | 0.51 ± 0.13 | 0.49 ± 0.28 | 0.76 ± 0.09 | 0.54 ± 0.14 |
| | NPV | 0.78 ± 0.06 | 0.81 ± 0.06 | 0.72 ± 0.02 | 0.79 ± 0.05 | 0.83 ± 0.05 |
| STACKING | AUC | 0.81 ± 0.05 | 0.72 ± 0.09 | 0.70 ± 0.10 | 0.83 ± 0.06 | 0.74 ± 0.10 |
| | F1 | 0.72 ± 0.06 | 0.50 ± 0.11 | 0.09 ± 0.12 | 0.73 ± 0.06 | 0.51 ± 0.11 |

| | | | | | | |
|---|---|---|---|---|---|---|
| | MCC | 0.50 ± 0.11 | 0.29 ± 0.16 | 0.03 ± 0.16 | 0.53 ± 0.11 | 0.34 ± 0.16 |
| | Sens. | 0.71 ± 0.07 | 0.52 ± 0.12 | 0.06 ± 0.08 | 0.72 ± 0.07 | 0.53 ± 0.15 |
| | Spec. | 0.78 ± 0.08 | 0.78 ± 0.08 | 0.95 ± 0.06 | 0.80 ± 0.09 | 0.80 ± 0.09 |
| | PPV | 0.74 ± 0.07 | 0.50 ± 0.12 | 0.39 ± 0.40 | 0.76 ± 0.09 | 0.53 ± 0.15 |
| | NPV | 0.76 ± 0.05 | 0.80 ± 0.05 | 0.71 ± 0.02 | 0.78 ± 0.05 | 0.82 ± 0.05 |
| SVC | AUC | 0.81 ± 0.05 | 0.72 ± 0.08 | 0.67 ± 0.08 | 0.83 ± 0.06 | 0.73 ± 0.09 |
| | F1 | 0.70 ± 0.06 | 0.48 ± 0.10 | 0.14 ± 0.14 | 0.72 ± 0.09 | 0.50 ± 0.15 |
| | MCC | 0.48 ± 0.11 | 0.29 ± 0.14 | 0.10 ± 0.17 | 0.53 ± 0.14 | 0.34 ± 0.20 |
| | Sens. | 0.67 ± 0.08 | 0.47 ± 0.11 | 0.09 ± 0.09 | 0.68 ± 0.12 | 0.49 ± 0.18 |
| | Spec. | 0.81 ± 0.07 | 0.81 ± 0.07 | 0.96 ± 0.06 | 0.83 ± 0.08 | 0.83 ± 0.08 |
| | PPV | 0.75 ± 0.07 | 0.51 ± 0.11 | 0.59 ± 0.36 | 0.77 ± 0.10 | 0.54 ± 0.19 |
| | NPV | 0.74 ± 0.05 | 0.79 ± 0.04 | 0.72 ± 0.02 | 0.77 ± 0.07 | 0.81 ± 0.06 |
| VOTING | AUC | 0.82 ± 0.05 | 0.73 ± 0.09 | 0.70 ± 0.07 | 0.83 ± 0.06 | 0.74 ± 0.09 |
| | F1 | 0.73 ± 0.07 | 0.52 ± 0.13 | 0.17 ± 0.14 | 0.74 ± 0.05 | 0.51 ± 0.12 |
| | MCC | 0.51 ± 0.12 | 0.32 ± 0.18 | 0.08 ± 0.15 | 0.53 ± 0.10 | 0.33 ± 0.15 |
| | Sens. | 0.72 ± 0.09 | 0.54 ± 0.15 | 0.12 ± 0.10 | 0.73 ± 0.07 | 0.53 ± 0.18 |
| | Spec. | 0.79 ± 0.08 | 0.79 ± 0.08 | 0.93 ± 0.05 | 0.80 ± 0.08 | 0.80 ± 0.08 |
| | PPV | 0.75 ± 0.07 | 0.51 ± 0.13 | 0.42 ± 0.27 | 0.76 ± 0.08 | 0.52 ± 0.14 |
| | NPV | 0.77 ± 0.06 | 0.81 ± 0.06 | 0.72 ± 0.02 | 0.79 ± 0.05 | 0.82 ± 0.05 |
| XGB | AUC | 0.79 ± 0.05 | 0.70 ± 0.08 | 0.69 ± 0.08 | 0.80 ± 0.06 | 0.72 ± 0.10 |
| | F1 | 0.70 ± 0.06 | 0.49 ± 0.12 | 0.06 ± 0.10 | 0.71 ± 0.06 | 0.49 ± 0.14 |
| | MCC | 0.47 ± 0.10 | 0.28 ± 0.17 | -0.01 ± 0.12 | 0.49 ± 0.12 | 0.29 ± 0.18 |
| | Sens. | 0.69 ± 0.08 | 0.51 ± 0.16 | 0.04 ± 0.08 | 0.70 ± 0.09 | 0.52 ± 0.18 |
| | Spec. | 0.77 ± 0.08 | 0.77 ± 0.08 | 0.95 ± 0.05 | 0.78 ± 0.10 | 0.78 ± 0.10 |
| | PPV | 0.73 ± 0.06 | 0.48 ± 0.11 | 0.26 ± 0.32 | 0.73 ± 0.09 | 0.48 ± 0.15 |
| | NPV | 0.75 ± 0.05 | 0.80 ± 0.05 | 0.71 ± 0.02 | 0.76 ± 0.05 | 0.81 ± 0.06 |

**Table S4. Comparative Performance of Machine Learning Models for Predicting Motor Fluctuations Onset.** The table displays key performance metrics (AUC, F1-score, MCC, Sensitivity, Specificity) under three distinct experimental conditions: models trained on all patients (first two columns), models directly trained only on patients without fluctuations at baseline (third column) and models trained using a random subset of patients matching in size the cohort of patients without motor fluctuations at baseline (columns four and five). Performance is evaluated on both the entire test set (All) and the clinically relevant subset of patients without motor fluctuations at baseline (Non fluctuating only). All values are presented as mean ± standard deviation. Sens.: Sensitivity, Spec.: Specificity, r.s.: random subset.

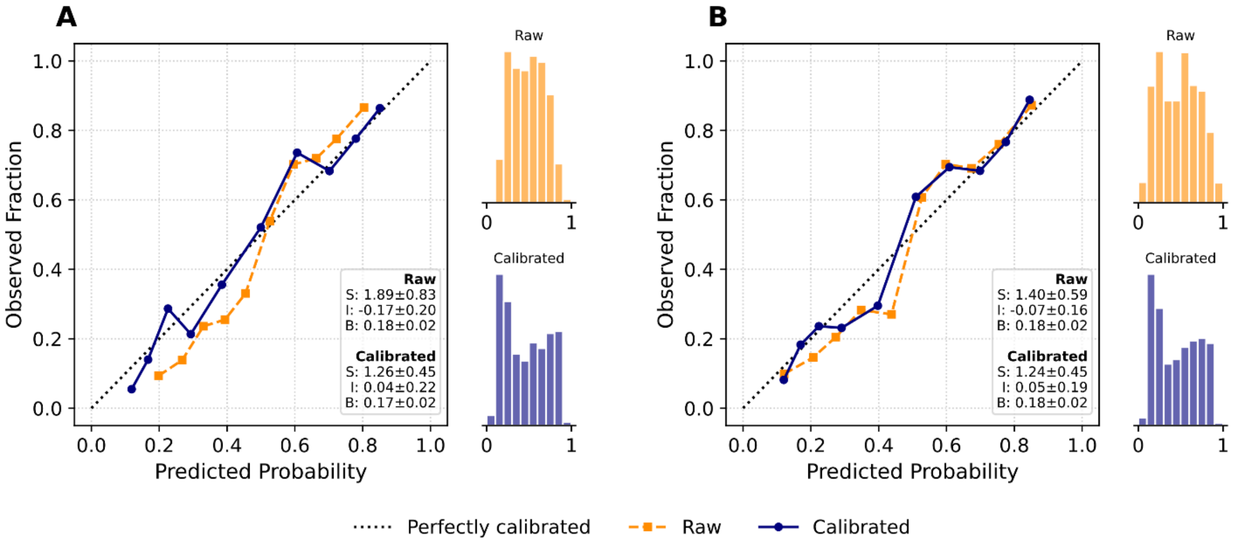


**Figure S3. Probability calibration analysis of the optimal predictive models - motor fluctuations task.** The plots illustrate the alignment between predicted probabilities (x-axis) and observed outcome frequencies (y-axis) for the two best-performing models: (A) Random Forest and (B) Voting ensemble. The orange squares (dashed line) represent the raw, uncalibrated classifier outputs, while the dark blue circles (solid line) depict the posterior probabilities after calibration via Platt Scaling. The diagonal dotted line indicates perfect calibration. Quantile binning (equal sample size per bin) was employed to ensure statistical robustness across the probability range. Inset text reports the quantitative calibration metrics: Slope (S, ideal=1), Intercept (I, ideal=0), and Brier Score (B, lower indicates better accuracy). Marginal histograms display the distribution of predicted probabilities, highlighting the shift from raw scores (orange) to calibrated probabilities (blue).

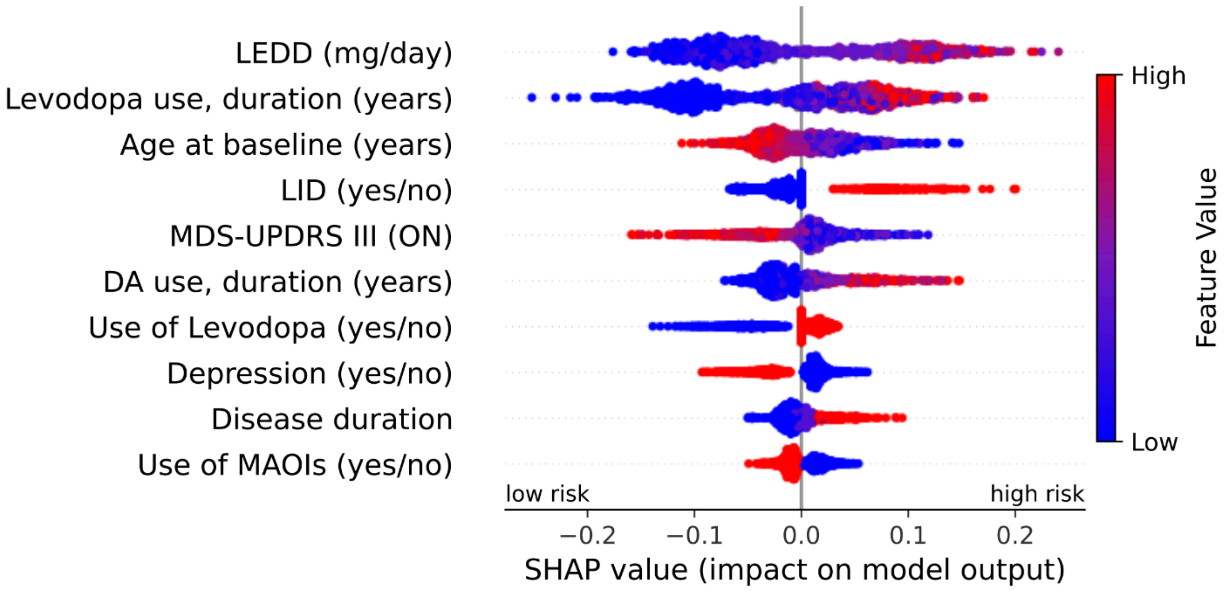


**Figure S4. SHAP analysis of Random Forest for the prediction of future motor fluctuations.** Global feature importance and impact for the RF model revealed by SHAP. Features are ranked in descending order of importance. Each dot represents a patient from the test set. The horizontal axis shows the SHAP value, indicating the feature's contribution to predicting the presence (positive values) or absence (negative values) of motor fluctuations within the third year. Color indicates the feature's original value (blue=low, red=high), revealing the direction and magnitude of its effect. Legend: DA: dopamine agonist; LEDD: levodopa equivalent daily dose; LID: Levodopa-induced dyskinesia; MAOIs: monoamine oxidase inhibitors.
